# A systematic review evaluating the effectiveness of virtual reality-based well-being interventions for stress reduction in young adults

**DOI:** 10.1101/2023.08.25.23294621

**Authors:** Joy Xu, Areej Khanotia, Shmuel Juni, Josephine Ku, Hana Sami, Vallen Lin, Roberta Walterson, Evelyn Payne, Helen Jo, Parmin Rahimpoor-Marnani

## Abstract

**Background:** Adolescents can be especially vulnerable to various stressors as they are still in their formative years and transitioning into adulthood. Hence, it is important for them to have effective stress management strategies. This systematic review investigates current well-being interventions that are aimed at reducing stress among young adults. In particular, interventions using the medium of virtual reality are explored.

**Methods:** This mixed-methods systematic review follows the PRISMA-P guidelines and articles were gathered for the databases PsycInfo, PubMed, Science Direct, Web of Science, Open Grey and Edutopia. Predetermined criteria and specific keywords were used to search for the articles. Search results were screened and extracted by two independent authors. Any disagreements after reconciliation were settled by a third author. The quality and risk of bias of included studies were assessed using the GRADE Quality Assessment Tool for Quantitative Studies. Studies were analyzed qualitatively.

**Results:** Among the appraised studies, the effectiveness of virtual reality-based interventions was measured in three contexts: nature, stress, and academics.

**Conclusion:** Studies using virtual reality interventions, overall, promoted a reduction in stress and an increase in well-being. The findings suggest VR may serve as an accessible and affordable medium of stress reduction for students and young adults. Larger sample sizes, and a greater number of included studies, may be required in future directions.

## INTRODUCTION

The COVID-19 pandemic has impacted millions of lives across the world and provided a spotlight on systemic disparities including present limitations to well-being and accessibility for support. The reverberations of this global phenomenon has reconciled greater focus on adolescents and young adults in particular, as their formative years were inevitably impacted by social distancing. The crucial years of their education were faced with obstacles in course delivery, academic opportunities, and social spheres. Furthermore, stresses were compounded with other hardships such as economic setbacks, limited socialization, and more issues that cumulatively burden one’s well-being, especially in the transition into adulthood. Thus, there has been significant effort to identify potential targets for interventions through research. Broadly, interventions aim to help study some change in individual experiences through strategies and processes after a systematic modification (APA, 2021). The main objective is to measure the effect of a process or program on certain situations (APA, 2021). In this review, virtual reality interventions refer to programs or treatments that target one or more determinants of health using a virtual reality (VR) headset, which displays a visual environment. This review refers to the Canadian Index’s definition of Well-being, stated as “The presence of the highest possible quality of life in its full breadth of expression focused on but not necessarily exclusive to good living standards, robust health, a sustainable environment, vital communities, an educated populace, balanced time use, …” (uWaterloo, n.d.). Thus, some aspects of an individual’s well-being can be measured through the extent to which an individual is stressed. Stress can be understood as a “psychophysiological response” to some form of danger, and involves biological components including nervous and hormonal responses to stimuli (Piotrowski and Hollar, 2019).

This systematic review questions the effectiveness of virtual reality interventions in reducing stress and promoting well-being in students and young adults. Currently, mindfulness-based interventions through virtual means are common, such as Mindfulness-Based Stress Reduction (MBSR), group mindfulness-based intervention (GMBI), and self-direct mindfulness-based intervention (SDMBI) through online delivery. There is evidence that these interventions have significant improvement in regulating emotion and mindfulness (Ma et al., 2018). However, there is little information regarding how virtual reality can be used for wellbeing interventions. Currently, the use of nature-based settings alone was found to improve mood to being “good” and “calm” in older adults, without the need for a mindfulness-based intervention curriculum (Kalantari, 2022). Thus, the articles included in our systematic review utilize auditory, visual, and even olfactory aids to simulate an MBI to reduce stress and increase mindfulness in young adults.

This systematic review aims to understand the effectiveness of current well-being virtual reality interventions in reducing stress among young adults. Due to the nature of learning in the 21st century relying heavily on online/virtual resources, implementing virtual tactics is imperative to combating these new sources of stress. In an era where generation Z is more stressed than previous generations and most are experiencing burnout (Carnegie, 2023), having virtual resources allows for immediate and low-maintenance aid. Felicity, as an application, strives to fill the need for these fast-acting resources via in-app modules, quizzes, and strategies.

## METHODS

A diverse range of literature, including grey literature was assessed according to inclusion/exclusion criteria, which was determined prior to the search. This criterion qualified the contextual and scientific relevance of the data by developing a standardized expectation for the literature’s content and experimental purpose. The PRISMA-P guideline was used for this qualitative review. Articles published between 1980-2022, that were randomized controlled trials, were gathered from PsycInfo, PubMed, Science Direct, Web of Science, Open Grey and Edutopia through an initial screening. A specific keyword search was used to gather these articles including the following keywords from each of the search platforms: (“wellbeing” OR “well-being” AND “student” AND “virtual” AND “reality” AND “quantitative” AND “intervention”). Duplicates and articles not matching the screening questionnaire were eliminated.

This review considers young adults to be within the age range of 15-40 years. All studies included are published in English. Studies included focused on the analysis of diverse student demographics, this was looked at in terms of; university programs, socioeconomic status, as well as culture, allowing for a robust analysis of student well-being. The interventions included can be applied virtually where concepts can be incorporated into a mobile application. Furthermore, stress reduction interventions that use either physiological or psychological questionnaires were included. Exclusion criteria included any research that generated qualitative data and studies published by organizations with conflicts of interest. Any research that does not have references available online, and studies with interventions geared towards individuals with psychological illnesses, and employs subjects with pre-existing psychological illnesses were also excluded.

Data for this study were collected and screened using the Covidence software. Multiple authors (AK, JK, HJ, LI, HS, EP) screened a collection of research for relevant studies in two steps: abstract screening and full-text screening. Two authors were assigned to each study. The authors worked independently and reconciled any disagreements and discrepancies after each step.

**Fig 1.**
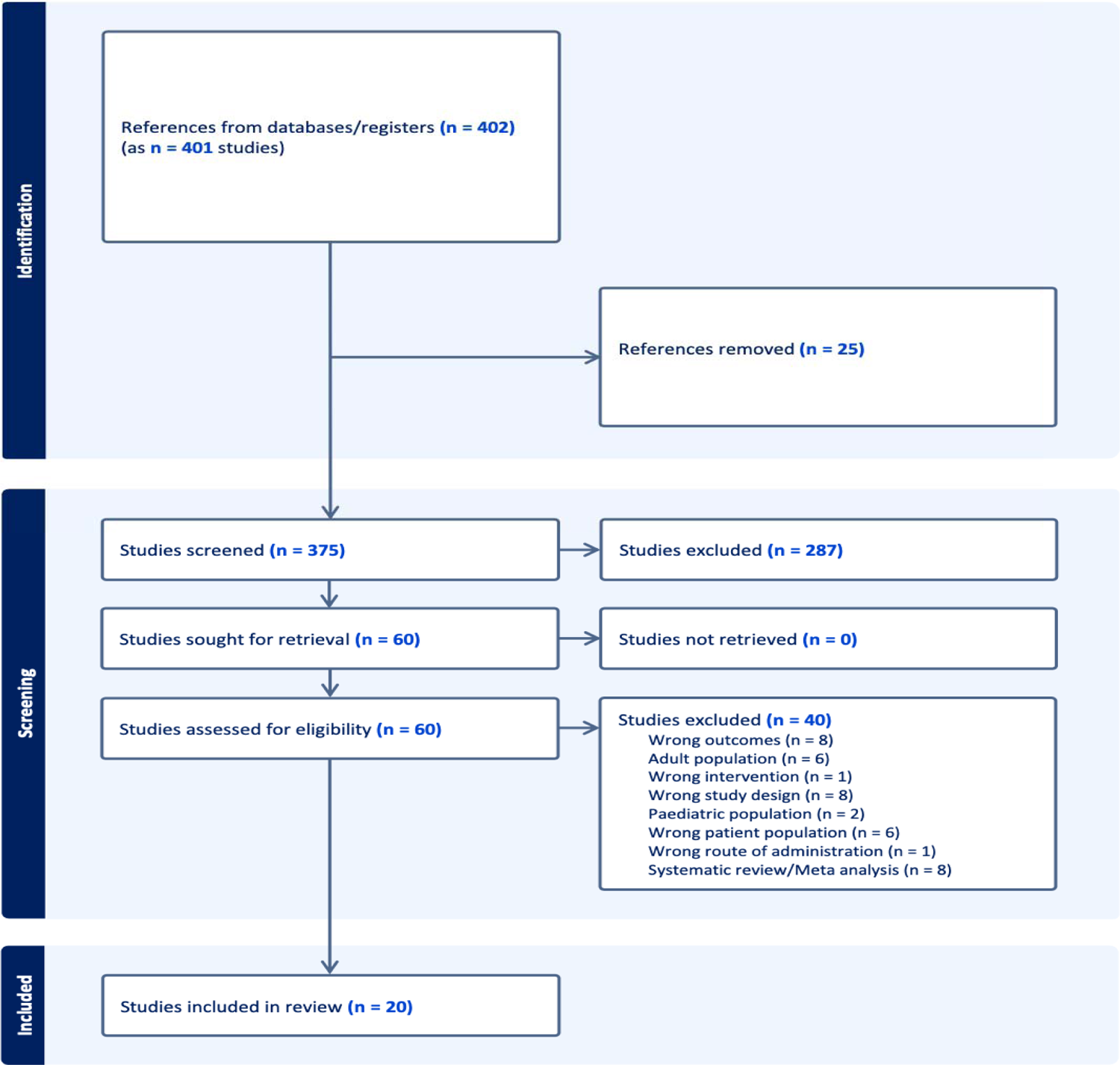
Structural diagram presenting screening and included studies

Following screening, the authors extracted relevant information to be used within this review, which included methods of study design, interventions, subject information, and outcomes.

Endnote was used to combine all studies found and to remove any duplicates. GRADE was used to assess the quality of the studies. GRADE consists of 4 levels of quality; very low, low, moderate, and high. The two authors independently used GRADE to assess the studies included and reconciled any disagreements coming to a final decision.

## RESULTS

**Table 1.1.** (Nature)

| Authors and year of publication | Methods | Population | Interventions | Main findings |
| --- | --- | --- | --- | --- |
| Deltcho Valtchanov and Colin Ellard, 2010 | Randomized control trial | Undergraduate students, aged 18-26, from the University of Waterloo, Canada.<br>Participants who can read/write English fluently, do not experience “seizures, vertigo, or motion sickness” or severe visual impairment. | 3 virtual reality settings:<br>1. Nature (tropical scenery)<br>2. Geometric (Assorted 3-dimensional shapes)<br>3. Urban (To-scale model of Shibuya station, Tokyo, Japan). | Mean positive affect scores in terms of ZIPERS scores were only significant in the Nature group post immersion in VR, with a mean of 2.21 (SD = 0.71) prior to VR, compared to 3.03 (SD = 0.98) post intervention ($p < 0.001$). Both geometric and urban VR settings had no significant impact on improving positive effect. |
| Allison O’Meara et al., 2020 | Randomized control trial | 1. 18 years old or older<br>2. Experience high test anxiety and no treatment<br>3. Control: Low or healthy level of test anxiety<br>4. From University College Cork, | 2 virtual reality settings:<br>1. Urban environment<br>2. Nature environment | Only the high anxiety group of students significantly benefited from the nature VR intervention within the 4 minute session ($p = 0.02$). The authors conclude that simulated exposure to nature can therefore reduce negative affect, thereby reducing test anxiety. Conversely, virtual |
|  |  | Ireland |  | exposure to nature settings did not significantly improve test scores. |
| Matthew H. E. M. Browning et al., 2020 | Randomized control trial | 18-27 years old, mean age 20.<br>University students, from the University of Illinois at Urbana-Champaign, USA<br>Excluded if: Diagnosed mood disorder, treated for mental illnesses, hearing impairments, use of alcohol/prescription drugs not normally taken in 24 hours. Intense physical activity in 24 | Control: Sit in front of blank wall<br>Outdoor group: Forest<br>Virtual Reality group: Same setting as forest group | Both VR and outdoor interventions have positive and significant results ($p < 0.001$), indicating high positive effect scores. All three conditions reveal significant decrease in negative affect values (outdoor group $p = 0.034$, control group $p < 0.001$, VR $p = 0.030$). Only the outdoor group reported statistically significant increase in positive effect for mood effects ($p = 0.044$). |
|  |  | hours |  |  |
| Tian Gao et al., 2019 | Randomized control trial | 120 Chinese college students, mean age 20.7.<br>Myopia degrees < 800. | 6 different VR environment interventions:<br>Grey (GrS)<br>Blue (BS)<br>Open green (OG)<br>Partly open green (POG)<br>Partly closed green (PCG)<br>Closed green (CG) | POG had the most impact on reducing negative mood, while CG had the least. Additionally, student preference for environments revealed that BS was most preferred, with GrS and CG as least preferred. A strong positive correlation is revealed between preference for a given environment, and positive mood improvement. |

**Table 1.2.** (Stress)

| Authors and year of publication | Methods | Population | Interventions | Main findings |
| --- | --- | --- | --- | --- |
| Daniela Villani & Giuseppe Riva. (2012) | Between-subject design | 36 Italian participants who were seen to be on or past the higher quartile on stress. | 1. 6 session stress management intervention<br>2. VR experimental: Used “ESCAPE” VR<br>3. Video experimental group<br>4. Audio experimental group | Participants were able to reduce their heart rate across conditions and with time. This was specifically seen in the VR condition, where they were more able to reduce their heart rate which in turn helped their emotional state. |
| Wayment, H. A., Collier, A. F., Birkett, M., Traustadóttir, T., & Till, R. E. (2015). | Randomized controlled trial | 32 female first-year undergraduate students | 1. Audio recording describing the 4 characteristics of the Quiet Ego.<br>2. Audio recording with VR of a park scene.<br>3. Control Group - read nature magazines | The VR with the audio Quiet Ego Contemplation reduced the degree to which participants felt “in the moment” as compared to the audio-only group ($p < 0.05$). |
| Villani, Daniela & Riva, Giuseppe. (2008). | Randomized controlled trial | 36 individuals who were students or office workers from Milan | 1. Nature VR (park, waterfall, river, garden etc.)<br>2. Nature Video (park, waterfall, river, garden etc.)<br>3. Relaxing Audio | VR, video, and audio of nature settings can help induce relaxation and help in stress management, increase positive emotions, enhance self-awareness, and contribute to emotional regulation. Of the three interventions, the VR intervention was seen to have the greatest psychological and physiological effects. |
| Villani, D.,<br>Riva, F., &<br>Riva, G.<br>(2007). | Randomized<br>controlled trial | 34 females and 30<br>males students<br>between the ages<br>of 21-28 years<br>from the Catholic<br>University of<br>Milan | 1. VR with video of<br>tropical islands<br>paired with audio<br>of therapeutic<br>sounds<br>2. DVD of relaxing<br>tropical videos<br>3. Audio of<br>therapeutic<br>narrative | VR of a relaxing nature<br>environment can<br>enhance positive<br>emotional states and<br>reduce anxiety. |
| Atul Taneja,<br>Vishal.S.B,<br>Mahesh.V,<br>Geethanjali.<br>B (2017) |  | Participants who<br>scored 14 or less<br>on the DASS 21<br>test were deemed<br>normal and<br>eligible for the<br>control group.<br>Participants who<br>scored 14 or more<br>were eligible for<br>the stress group. | 1. STROOP task<br>2. VR-based stress<br>therapy<br>intervention | PANAS questionnaire:<br>mean score of 32 for<br>positive affect and a<br>mean score of 15 for<br>negative affect were<br>generated |

**Table 1.3.**
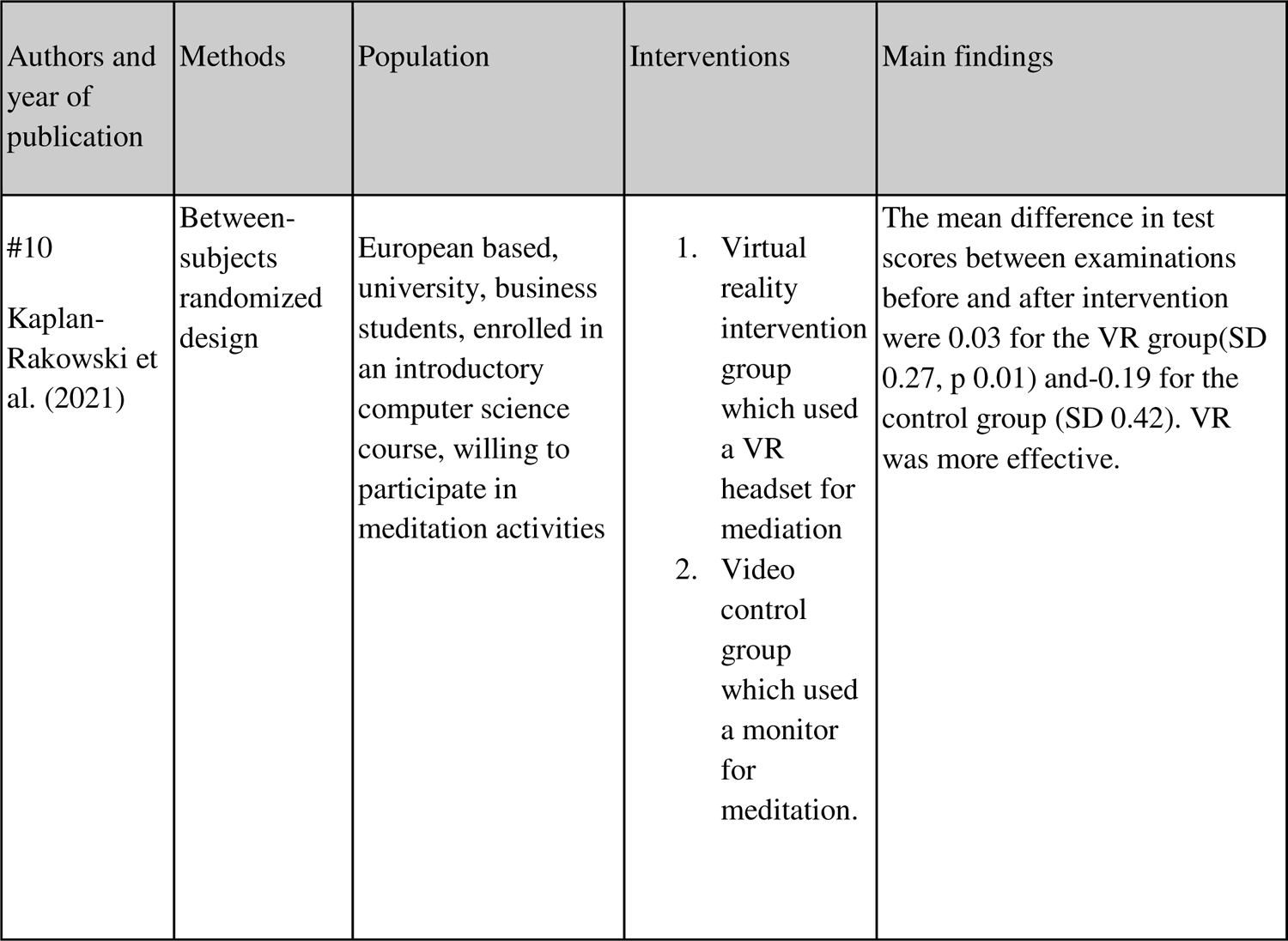
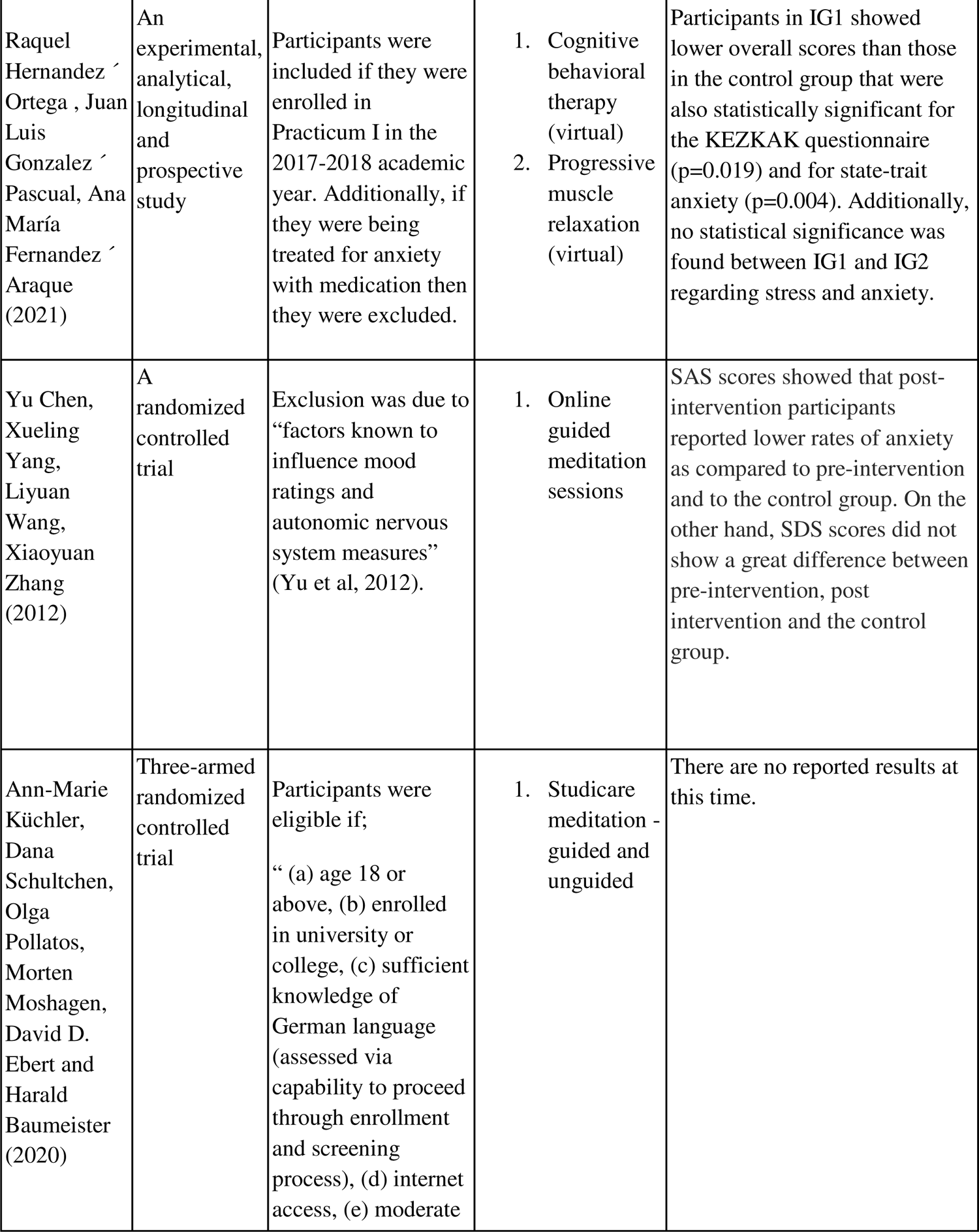
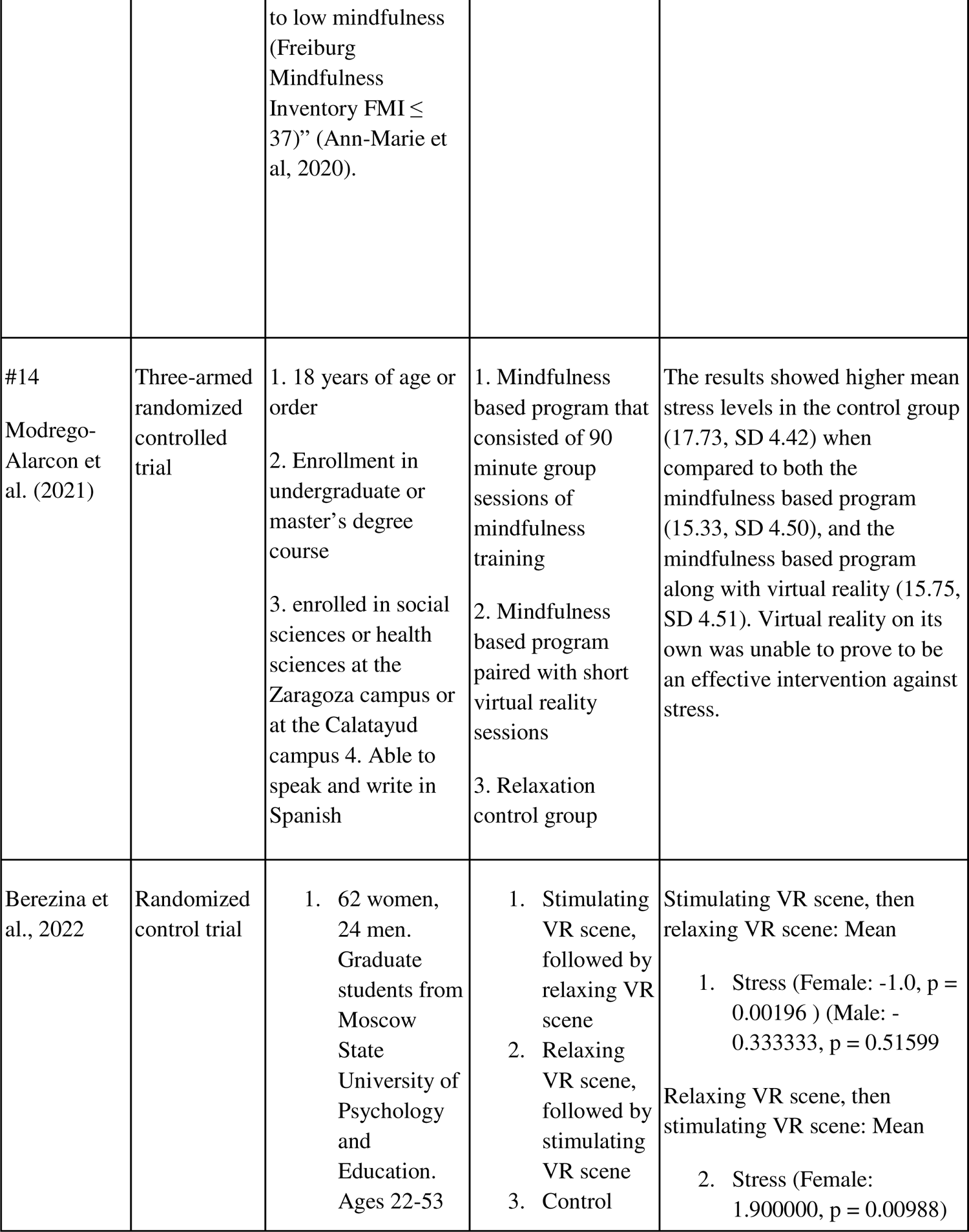
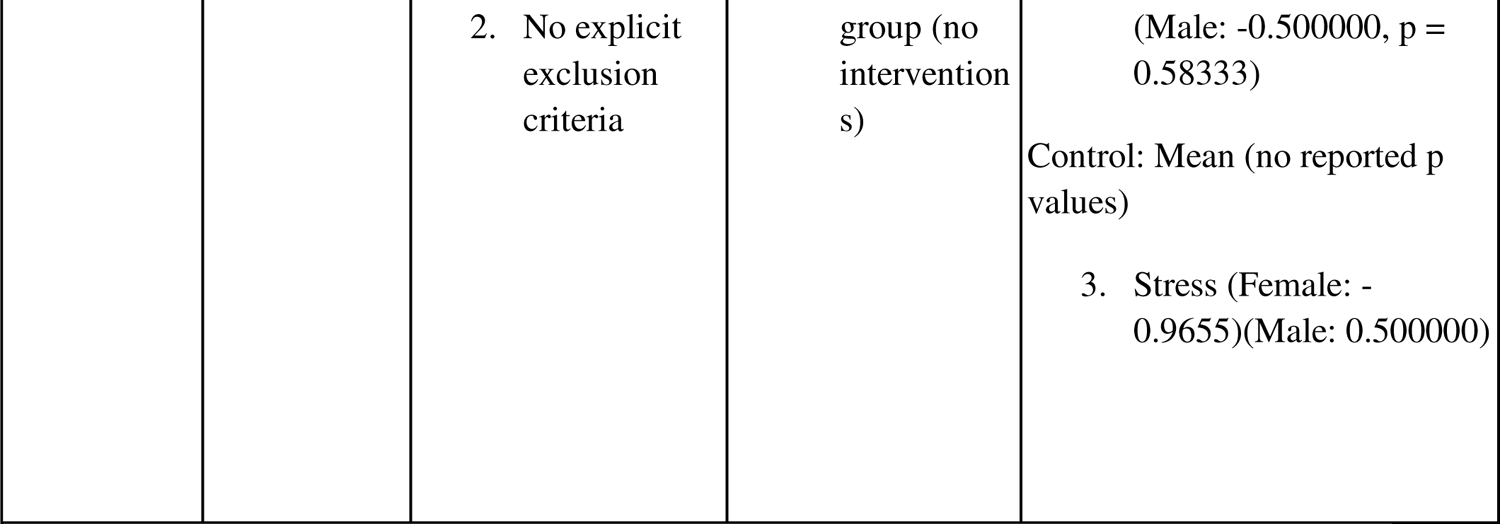
(Academic Contexts)

## DISCUSSION

### Measures of wellbeing through exposure to nature

From the included studies that underwent data extraction, a considerable number explored the role of natural environments in a virtual reality (VR) setting on wellbeing. It is well known that exposure to nature can positively improve wellbeing, such as daily walks in urban parks, hikes, or gardening (Weir, 2020). In a study by White et al. (2019), spending two hours in nature per week revealed odds ratios for good health and high well-being that were significantly higher than the control of no nature exposure, at 1.20 and 1.33 respectively, compared to 1.0 of the control.

However, access to stimulating biodiversity may not be possible for every individual, such as students in highly-urbanized campuses, (dis)abled individuals, and students who live in climates with long winter seasons. Therefore, virtual reality may serve as an accommodation. Browning et al. (2020) reveal that 6 minutes of VR in an outdoor nature setting results in high positive affect levels post-intervention (p<0.001). The study emphasizes the importance of 360-degree nature videos that are immersive and interactive, for adequate similarity to the natural world. O’Meara et al. (2020) revealed that students with high anxiety can benefit from VR nature exposure, through significantly reduced negative affect (p = 0.02). The authors argue that these negative effects can be associated with exam anxiety, thus VR serves as a well-being tool to reduce student stress.

The significance of the setting in the VR experience, namely nature, over other landscapes is explored within Valtchanov’s study. A neutral geometric environment, an urban environment, and a nature environment were randomly allocated to students with induced stress. The nature setting increased positive effects significantly (p<0.001) from a mean of 2.21 pre VR to 3.03 post-VR, compared to no effect of the other settings. Self-reported stress significantly decreased in the nature environment (p<0.005), unlike the geometric or urban settings. Additionally, differences in the biodiversity of natural environments in VR were also compared within the included studies. Gao et al. (2019) conclude that, despite no significant differences between the VR settings regarding impact on physiological stress, positive mood, or attention, the authors reveal that restorations in negative mood were significantly different (p = 0.03). Partly open green spaces, defined in the study as having a 10-30% composition of trees and shrubbery in the VR environment, had the highest significant effect on reducing negative mood, at p<0.01. To summarize, natural environments presented in the medium of VR can promote positive mood, lowered stress, and overall well-being that can supplement a lack of access that students may face. The included studies feature VR through headsets, such as the Oculus Rift. While immersive, these devices are expensive and inaccessible to the average student. Thus, future directions of implementing VR through cell phones that can mount onto cheap headsets, such as the Google Cardboard paired with audio, may be a cost-efficient but impactful VR intervention with nature.

### Measures of well-being via stress levels

Five of the included studies discussed interventions influencing well being as seen through measures of stress levels. Among many therapy approaches to reduction of stress and anxiety, relaxation techniques are seen to be very effective (Atul et al, 2017). Atul et al (2017) explored this by demonstrating the effectiveness of virtual reality therapy through the use of virtual reality environments and relaxing audio. Through the use of Positive and Negative Affect schedule (PANSAS) questionnaire, a mean score of 32 for positive affect was found as compared to a mean score of 15 for negative affect. Similarly, a study by Plante et al (2003) looked at the impacts of virtual reality (paired with aerobic exercise in a laboratory setting) on well-being and stress. Stress and energy were measured under four conditions: exercise outdoors, exercise with virtual reality in laboratory conditions, exercise without virtual reality in laboratory conditions and virtual reality without exercise. Outcomes were measured using AD-ACL checklists. It was found that exercising outdoors resulted in the most significant decreases in stress and increases in energy in men (p<0.1) and especially in women (p< 0.5). Women also felt calmer (p<0.5) following the VR intervention, though the findings were not significant for the men. While there is room for more research, this finding strengthens the credibility of virtual reality as an intervention to reduce stress.

The rapid development of technology in recent years has impacted how one perceive, interprets and organizes their day-to-day lives, which may influence one’s health and well-being (Zhou et al., 2020). When assessing well-being, an aspect that may be considered includes an individual’s sense of presence, which can be described as how one experiences and engages in events and situations at the moment (Zhou et al., 2020). Specifically, in the case of virtual environments and simulations, presence can be referred to as an experience where the user is immersed in the VR environment (Grassini & Laumann, 2020). A study conducted by Villani et. al. (2007) found that VR intervention involving an environment with nature can help induce psychological and physiological effects such as relaxation (p < 0.005) and lowered anxiety (p < 0.01). A similar study conducted by Villani, D., & Riva, G. (2008) reported that VR can significantly reduce anxiety levels (p < 0.05), manage a state of anxiousness (p < 0.005), and increase relaxation (p <0.01). Both studies the State-Trait Anxiety Inventory (STAI) to measure anxiety levels along with the Coping Orientation to Problems Experienced Questionnaire (COPE) Questionnaire to assess the stress levels of participants (Villani et. al., 2007;Villani, D., & Riva, G., 2008). In the past, researchers have suggested that a greater sense of presence can increase the level of engagement with a virtual simulation (Cummings & Bailenson, 2016; Grassini & Laumann, 2020). One study conducted by Mostajeran et al. (2023) assessed the cognition and stress levels of office workers before and after a VR intervention involving virtual plants. It was found that a virtual environment with plants (SD = 0.82) significantly increased the sense of presence (p < 0.05), compared to a virtual environment where there were no plants (SD = 0.87) (Mostajeran et al., 2023). Additionally, it was observed that participants performed significantly better on memory tests such as the digit span backward test (p < 0.05) after the VR intervention (Mostajeran et al., 2023). The findings by Mostajeran et al. (2023) suggest that nature VR environments can help promote a sense of presence, which can have positive effects on productivity levels, as well as mediating stress, which aligns with the findings in the current study.

However, not all of the studies showed a benefit from VR. In a study from 2015, 32 female undergraduate students dealing with stress and anxiety in their transition to university completed a quiet ego contemplation intervention, which reminded them of four characteristics of the quiet ego: detached awareness, inclusive identity, perspective taking, and growth (Wayment et al., 2015). They were split into 3 groups: an audio recording describing the four characteristics, the same audio recording paired with virtual reality of a park scene, and a control group in which participants perused nature magazines. The participants completed three 15-minute sessions of their assigned condition over 6 weeks. The study found that adding the VR component to quiet ego contemplation reduced the degree to which participants felt “in the moment” (*t*(28)=2.66, *p*<0.05). The authors hypothesized that this could be explained by the low quality headset that was used, which was uncomfortable for participants, and by the self-directed aspect of the VR, which may have been more distracting than a guided VR experience would have been.

### Measures of well-being via academia contexts

The final theme extracted from the studies included covered measures of well-being as seen in academic contexts. These studies included participants in academic institutions who are facing many stressors as a result of their academic career. Two of those studies recruited nursing students; one study utilized cognitive behavioral therapy and progressive muscle relaxation (Ortega et al, 2021), and the other study administered a mindfulness meditation intervention (Chen et al, 2013). Ortega et al (2021) used the KEZKAK questionnaire and the state-trait anxiety test as a measuring tool for stress levels. They found overall lower scores for both tests amongst participants who underwent the intervention with p-values of 0.019 for the KEZKAK questionnaire and 0.004 for the state-trait anxiety test (Ortega et al, 2021). Similarly, Chen et al (2013) used the self-rating anxiety scale as a measurement tool where lower rates of anxiety were reported for participants in the intervention group. In considering these findings, the strength of virtual-based interventions to decrease stress is further reinforced. The authors of both studies argue that these types of intervention are necessary for students to be able to function healthily and excel in their fields (Ortega et al, 2021; Chen et al, 2013).

A common intervention method among these studies was the use of VR headsets. Kaplan-Ratkowski, Johnson and Wojdynski (2021) utilized VR headsets in their study for a meditation intervention. This was used for the experimental group, while the control group went through the intervention by watching it on a monitor. Participants went through a pretest and posttest which entailed basic computer science tasks. It was found that the VR group (0.03) showed a higher mean difference in test scores as compared to the control group (−0.19). Berezina et al’s (2022) intervention also used a VR headset. Participants in experimental group A watched a stimulating scene, followed by a relaxing scene, while participants in experimental group B watched a relaxing scene followed by a stimulating scene. Finally, there was a control group, which did not watch anything. They found that while both experimental groups experienced a decrease in fatigue, experimental group B showed a less pronounced decrease as compared to experimental group A. The final study in this theme utilized a virtual reality intervention aimed at reducing stress. There were three groups, one which underwent a group mindfulness session, another which went through a virtual reality session, and a control group. Modrego-Alarcon et al (2021) found that participants in the mindfulness-based program showed the lowest levels of stress as compared to any other group. Additionally, the virtual reality group scored lower stress levels as compared to the control control group. Overall, the studies show that virtual reality, well-being interventions are effective in reducing stress levels and promoting relaxation among students, especially those in higher institutions.

## CONCLUSION

In conclusion, this systematic review aimed at evaluating the efficacy of virtual reality interventions to promote the well-being of students and young adults. To achieve this, studies were divided into themes of nature, stress, and academic contexts as focuses of interventions. Overall, the included studies reveal that virtual reality interventions pose as a promising medium to reduce the stress experienced by young adults and students, which can ultimately improve wellbeing. These findings reveal that VR may serve as an accessible, affordable tool for students and young adults to promote well-being/lower stress levels. However, there are some limitations to the review. The included studies tend to have smaller sample sizes, which may not be representative of students as a whole. To add, only 20 studies were included in the final phase of extraction. Future directions may include expanding the search criteria to include more studies that may have higher sample sizes.

## Data Availability

All data produced in the present study are available upon reasonable request to the authors

## Acknowledgements

Liza Iaralova, Valerie Lo, Cherry Tagra, Ammal Riaz

## ABBREVIATIONS

VR: Virtual Reality

